# Changes in Work-Life Balance Experienced Amongst Ophthalmologists During the COVID-19 Pandemic

**DOI:** 10.1101/2025.08.27.25334594

**Authors:** Rucha K. Borkhetaria, Mckenzee Chiam, Erik Lehman, Amanda L. Ely

## Abstract

**Purpose:** To evaluate professional versus non-professional ophthalmologist responsibilities and how this changed through the COVID-19 pandemic.

**Methods:** American Academy of Ophthalmology members were emailed a survey between December 2020-January 2021. The percentage of time spent on professional, childcare, household responsibilities pre- and amid-pandemic (after March 11, 2020) were evaluated, along with professional and non-professional stressors.

**Results:** Of 1,250 ophthalmologists, 150(12%) responded, 43%(64/150) women and 56%(84/150) men. Pre-pandemic, fewer women worked full-time compared to men [83%(53/64) vs. 95%(80/84); p=0.013]. Men reported a decrease in work hours from pre- to amid-pandemic (46.4 vs. 44.6hours; p=0.010); women reported no significant changes (44.2 vs. 44.5hours; p=0.699). Women spent an increased amount of time from pre- to amid-pandemic on overall household (18.4% vs. 21.8%, p<0.001) and childcare responsibilities (18.5% vs. 21.1%, p=0.014). This was not seen amongst men pre- to amid-pandemic [household: (21.2% vs. 22.4%, p=0.057); childcare: (9.2% vs. 9.8%, p=0.104). Overall stress levels increased amid-pandemic (31.5% to 43.1%, p<0.001), seen amongst men (31% to 39.1%, p<0.001), and women (32.5% to 47.9%, p<0.001).

**Discussion:** Increased overall stress was seen in ophthalmologists as a result of the COVID-19 pandemic. Further investigation is required to understand the relationship between the pandemic and ophthalmologists’ burn-out.

## INTRODUCTION

The proportion of women entering medicine has dramatically increased in recent decades. According to the Association of American Medical Colleges (AAMC), women accounted for 34.0% of medical school graduates in 1990, compared to 47.9% of the graduates in 2019.^1^ Despite this increase in representation of women in medicine, women remain underrepresented in many surgical specialties and remain concentrated in pediatrics, internal medicine, and family medicine.^2^ In 2019, while women represented 36.3% of practicing physicians across all specialties, women represented approximately one quarter (26.7%) of all practicing ophthalmologists.^3^ Although many factors may influence one’s specialty choice, West et al. reported that lifestyle and the amount of time available to spend with family were the main factors in career decision-making amongst women.^4^

Pre-pandemic studies have shown gender differences in how the current generation of physicians allocate time for professional versus parental/domestic work.^2,5^ A prior study by Jolly et al. showed that among married physicians with children, women reported spending greater than eight hours per week more on domestic activities compared to their male colleagues.^5^ There is a lack of research, however, evaluating whether or not these gender differences in parental/domestic versus professional responsibilities carry over into ophthalmology.

The impact of the Coronavirus Disease 2019 (COVID-19) on professional and non-professional responsibilities is vast. The COVID-19 pandemic has presented unparalleled challenges in the healthcare landscape, both professionally and non-professionally, with increases in work demands, personal health risks of contracting COVID-19, and domestic and childcare responsibilities.^6^ Prior studies have suggested that women are disproportionately affected during times of disaster due to increases in productive, reproductive, and community responsibilities.^7^ Already, the COVID-19 pandemic has amplified pre-existing inequities affecting women in all fields of medicine, including ophthalmology^8^, submitting fewer publications compared to their male colleagues during the pandemic.^9^

In this study, we aimed to investigate gender differences in domestic responsibilities and stressors during both pre-pandemic and amid-pandemic times in ophthalmology, to pave the way for targeted interventions to equalize success and promote wellbeing.

## METHODS

This cross-sectional survey study was approved by the Institutional Review Board of Penn State College of Medicine, adhered to the tenets of the Declaration of Helsinki and its later amendments, and maintained Health Insurance Portability and Accessibility Act (HIPAA) compliance. Completion of the survey was considered implied consent to participate in the study.

American Academy of Ophthalmology members who were active in 2020 (1250 members total) were invited by email to participate in a confidential, 59 question online survey. Data on gender, ophthalmologic specialty, marital status, family size, professional responsibilities, and division of childcare and household responsibilities, both pre- and amid- the COVID-19 pandemic were collected. The pre-pandemic period was defined as before March 11, 2020, and the amid-pandemic period between March 11, 2020 and the survey collection date of January 2021. Questions were scored using a 5-point Likert scale. Perceived stress was measured using a sliding scale of 0-100% with 0% being no stress perceived and 100% being greatest amount of stress perceived. Survey results were collected using REDCap, an encrypted web-based database, hosted at Penn State College of Medicine. A reminder notice was issued 2, 3, and 4 weeks after the initial survey invitation.

All variables were summarized prior to analysis, and the distributions of continuous variables were assessed using histograms, normal probability plots, and tests for normality to aid in determining what type of tests were appropriate. Chi-square tests, two-sample t-tests, and Wilcoxon Rank-Sum tests were conducted to analyze gender differences within pre-pandemic and amid-pandemic time points. McNemar’s tests, paired t-tests, and Wilcoxon Signed Rank tests were used to test for differences between the time points overall or within genders. Quantile regression of the median tested for differences between genders in the change in outcomes from pre-pandemic to amid-pandemic. SAS version 9.4 (SAS Institute, Cary, NC) was employed to perform all analyses, and all comparisons were made with a significance level of 0.05.

## RESULTS

### Demographics

Of 1,250 ophthalmologists invited to participate in this study, 150 responded (12%), of whom 64 (43.0%) were women and 84 (56.0%) were men. Table 1 depicts the general characteristics of respondents.

**Table 1:** Study Population Characteristics.

| Ophthalmologist Characteristics | Total | Female | Male | P-value |
| --- | --- | --- | --- | --- |
| *n* (%) | 150 | 64 (43) | 84 (56) |  |
| Age, mean (SD) | 53.8 (10.7) | 49.3 (9.2) | 57.5 (10.5) | <0.001 |
| Ophthalmology Work Hours, mean (SD) |  |  |  |  |
| Before Pandemic | 45.3 (13.8) | 44.2 (14.3) | 46.4 (13.5) | 0.298 |
| During Pandemic | 44.4 (15.3) | 44.2 (15.2) | 44.4 (15.4) | 0.944 |
| Have Children, *n* (%) | 140 (93.9) | 58 (95.6) | 81 (98) | 0.490 |
| Number of Children, mean (SD) | 2.4 (1.1) | 2.2 (0.9) | 2.5 (1.2) | 0.077 |
| Child with Disabilities, *n* (%) | 6 (4.3) | 2 (3.5) | 4 (4.9) | 0.705 |
| Partner Profession Before Pandemic, *n* (%) |  |  |  |  |
| Unemployed | 10 (7.6) | 1 (0.8) | 9 (6.8) | 0.028 |
| Stay-at-home parent | 20 (15.2) | 5 (3.8) | 15 (11.4) | 0.028 |
| Employed | 103 (77.3) | 47 (35.6) | 55 (41.7) | 0.028 |
| Partner Profession Amid-Pandemic, *n* (%) |  |  |  |  |
| Unemployed | 7 (7.3) | 0 (0) | 7 (100) | 0.015 |
| Employed | 89 (92.7) | 43 (44.8) | 46 (47.9) | 0.015 |

Note: Respondents who answered "Rather not say" or "Other" to the survey question asking for gender were excluded from the analysis.

Since one respondent answered “rather not say” to the survey question asking for gender, and one did not answer the question, they were removed from our binary gender analyses. Our study included 133 (89.9%) full-time ophthalmologists [95.2% of males reported working full time vs. 82.8% of females (*p*=0.013)], and 15 (10.4%) part-time ophthalmologists, [4.8% of males reported working part-time vs. 17.2% of females (*p*=0.013)]. Subspecialties of oculoplastics and refractive surgery had significantly more men than women, while pediatrics showed significantly more women than men (Figure 1).

**Figure 1.**
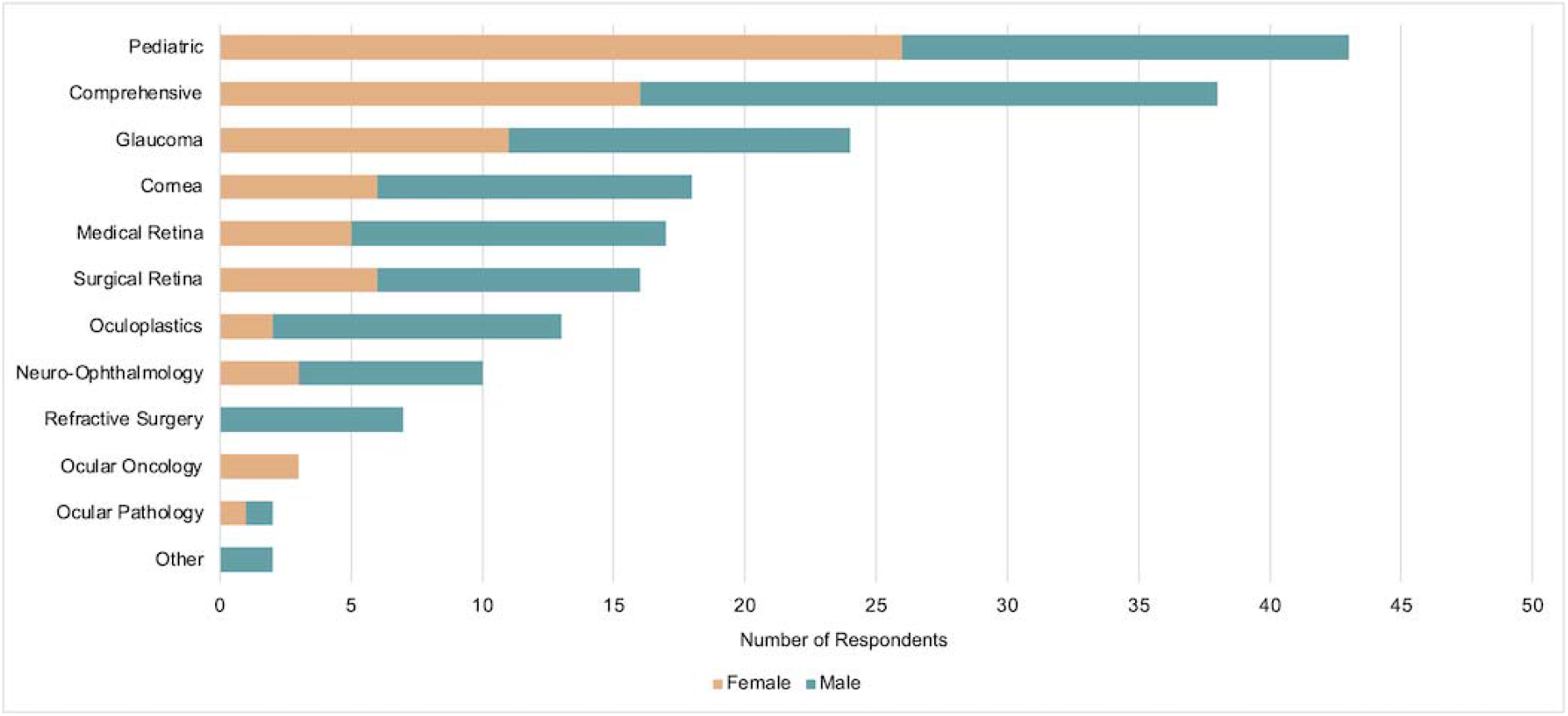
Ophthalmology Subspecialty Demographics. **\***: p<0.05

Of the 132 ophthalmologists who were married or in a domestic partnership, 20 (15.2%) reported that their partner was a stay-at-home parent before the pandemic, more commonly reported amongst male compared to female ophthalmologists [15/79 (19.0%) vs. (5/53 (9.4%); p=0.028]. Among those respondents whose spouses had a job pre-pandemic, 19.6% (21/107) of respondents reported their spouses’ jobs were affected by the pandemic, whereas 80.4% (86/107) reported their spouses’ jobs remained the same during the pandemic. No gender difference was noted in these analyses (p=0.920).

### Professional Responsibilities

Female and male ophthalmologists reported similar work-hours pre-pandemic (44.0 vs. 46.4 hours/week; p=0.298) and amid-pandemic (44.2 vs. 44.4 hours/week; p=0.944). While female ophthalmologists did not report a significant change in work hours pre- vs. amid-pandemic (44.2 vs. 44.6 hours/week; p=0.699), male ophthalmologists reported a significant decrease in work hours of approximately two hours/week (46.4 vs. 44.6 hours/week; p=0.010).

### Domestic Responsibilities: Household

Figure 2 presents a multivariate model of time spent on household and childcare responsibilities by gender and pandemic status. Overall, ophthalmologists reported spending an increased amount of time pre- to amid-pandemic on overall household responsibilities (19.9% pre- to 22.1% amid-pandemic; p<0.001). When analyzing for a change from pre- to amid-pandemic by gender, women reported spending an increased amount of time on overall household responsibilities (18.4% pre- to 21.8% amid-pandemic, p<0.001); this was not seen amongst men (21.2% pre- to 22.4% amid-pandemic, p=0.057).

**Figure 2.**
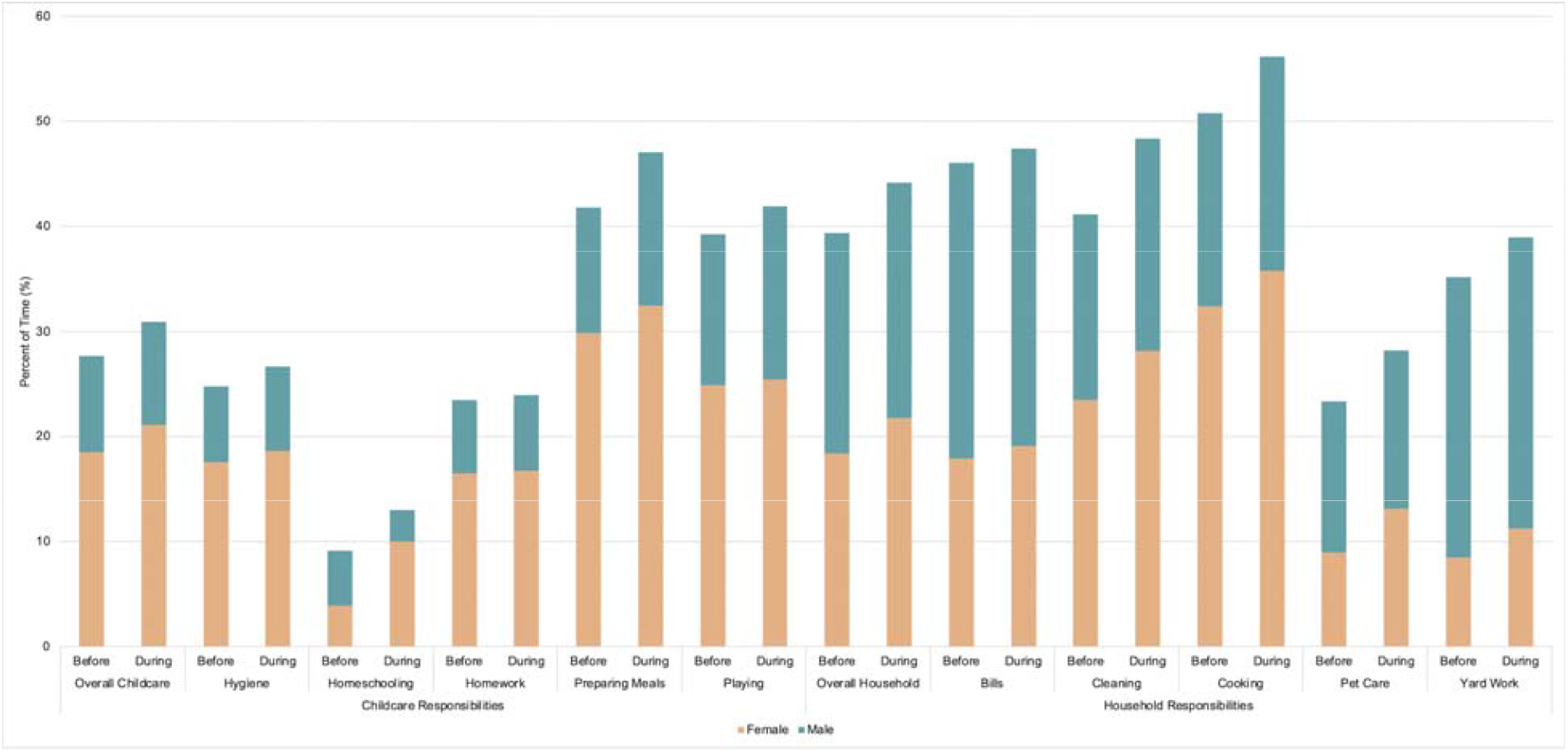
Percent of Time Female and Male Ophthalmologists Spent on Childcare and Household Responsibilities Before and During the COVID-19 Pandemic **\***: p<0.05

### Domestic Responsibilities: Childcare

This analysis was restricted to those ophthalmologists who reported having children. The majority of ophthalmologists reported having at least one child (93.9%), with 60.0% reporting that their child/children lived at home with them. Ophthalmologists with children reported spending an increased amount of time on overall childcare responsibilities pre- to amid-pandemic (13.3% pre- to 14.8% amid-pandemic; p=0.002). Further gender analysis revealed that women reported a significant increased amount of time pre- to amid-pandemic on overall childcare tasks (18.5% pre- to 21.1% amid-pandemic, p=0.014); this increase was not significant among men (9.2% pre- to 9.8% amid-pandemic, p=0.104). See Figure 2.

### Stress Level

Figure 3 presents a multivariate model of self-reported stress levels by gender and pandemic status. Women and men reported similar overall stress levels pre-pandemic (women 32.5% vs. men 31.0%, p=0.821); however, amid-pandemic, women reported a greater overall stress level compared to men (women 47.9% vs. men 39.1%, p=0.012). Both women and men reported an increase in overall stress pre- to amid-pandemic [women: increase of 15.4% (32.5% to 47.9% p<0.001), men: increase of 8.1% (31.0% to 39.1%, p<0.001)], with women showing a significantly greater increase in overall stress amidst the pandemic when compared to men (p<0.001). Overall stress levels before the pandemic were similar between ophthalmologists whose children lived with them at home and ophthalmologists whose children did not (33.8% vs. 30.3%, p=0.181); amidst the pandemic, however, those with children at home reported greater overall stress levels than those without children at home (47.7% vs. 39.7%, p=0.013).

**Figure 3.**
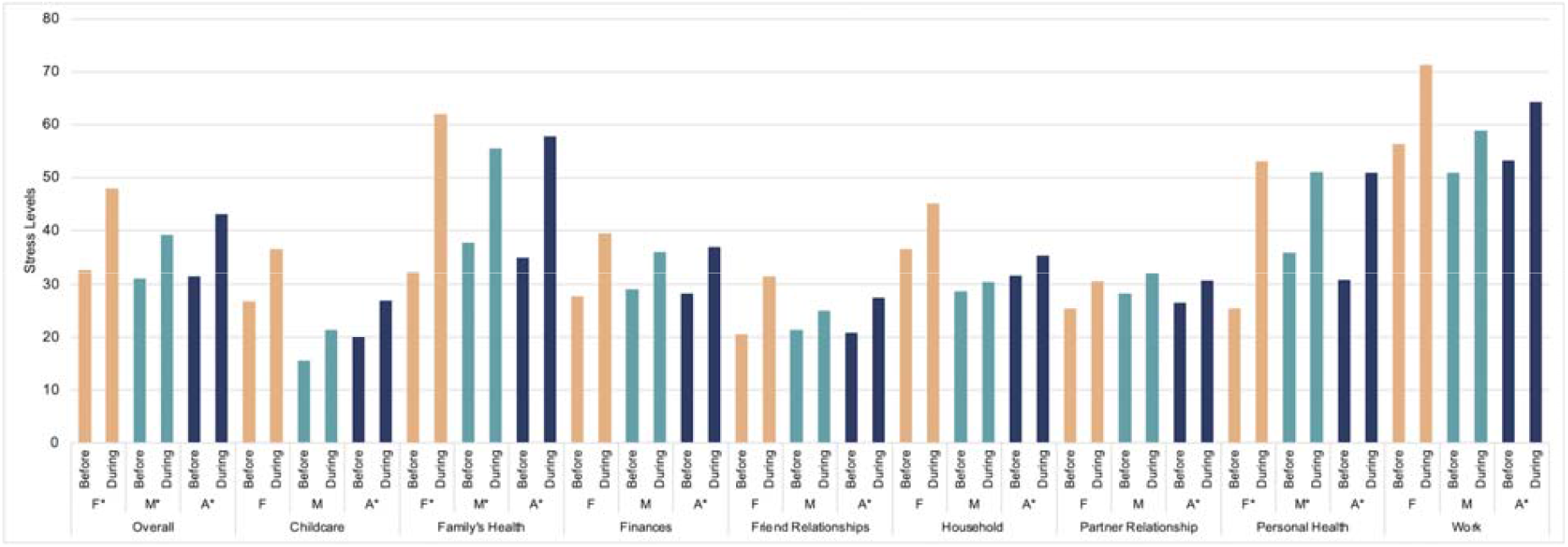
Ophthalmologist Stress Levels According to Gender Before and During the COVID-19 Pandemic **F:** Female **M**: Male **A**: All *: p<0.05

## DISCUSSION

In this national examination of ophthalmologists in the early period of the COVID-19 pandemic, we found that both female and male ophthalmologists experienced a rise in overall stress; female ophthalmologists, however, reported a disproportionately greater domestic and professional burden in terms of greater levels of overall stress and increased domestic responsibilities compared to their male colleagues.

We consulted the literature to investigate potential causes of greater stress amongst women compared to men in general during the COVID-19 pandemic. In the early period of the pandemic, governments worldwide implemented stringent guidelines to prevent uncontrolled spread of the virus, with up to 153 countries participating in school closures, affecting approximately 1.2 billion, or 68%, of students worldwide.^10^ Within the United States, up to 48.7 million people, or approximately 35% of the employed workforce, began to work from home in May of 2020.^11^ Literature has demonstrated that in pre-pandemic times, unpaid work such as childcare and housework increased women’s stress.^12^ School closures, work-from-home orders, and a decrease in outsourced childcare and household duties during the pandemic may have led to a greater domestic burden that may disproportionately affect women. Although dual-earner households are becoming more prevalent, women have been shown to still bear the greater responsibility for housework and childrearing^13–17^, even amongst physicians.^18,19^ Moreover, several studies reveal that the pandemic imposed a worsening domestic burden upon mothers compared to fathers^16,17,20,21^, seen amongst physicians as well.^22,23^

Ophthalmologists with children experienced an overall increased level of stress due to the pandemic compared to those without children. These same ophthalmologists reported an increase in the amount of time spent and stress related to their childcare responsibilities. As for household responsibilities an equal percentage of time spent pre-pandemic was reported for both female and male colleagues. Further gender analysis, however, revealed that from pre- to amidst the pandemic, only women reported an increased amount of time spent on childcare and household responsibilities associated with an overall higher level of stress compared to men in these categories regardless of pandemic status. Linzer et. al proposed that gendered expectations of female clinicians may contribute to both part-time work status and a higher rate of burnout among female physicians.^24^ Though we had a small sample size of part-time clinicians participating in this survey, within the ophthalmology community it appears that more female physicians may hold part-time job status when compared to their male colleagues. A concerning report from Japan stated that approximately 70% of female physicians leave work for childbirth and childrearing responsibilities.^25^ The significant burden of childcare and household responsibilities brought on by the COVID-19 pandemic likely contributed, in part, to the increase in overall stress reported by women ophthalmologists. However, the increased stress reported by women even prior to the pandemic in both childcare and household responsibilities, warrants further investigation in order to aide in the development of programs that will help alleviate stress to the physician-parent population as a whole.

There has been speculation that the increased domestic responsibilities women faced during the early months of the pandemic may have had an adverse effect on women’s work and work-related burnout.^26^ Further evidence suggests that female physicians may be experiencing greater rates of burnout during this pandemic.^27^ Although our study did not ask about ophthalmologist burn-out in particular pre- vs amid-pandemic, this area does warrant further investigation.

The effects of the pandemic have not only entered the home, but extended into professional life as well. COVID-19 has exacerbated the authorship gender gap^9,28^, even within ophthalmology^8^. In our study male ophthalmologists reported a significant decrease in work hours during the pandemic, while female ophthalmologists did not. Our study is limited in that we did not ask respondents to provide a reason for a decrease in work hours and thus we cannot interpret why men would lose more work hours compared to their female colleagues. Our study also did not look into the work-related stressors of telemedicine. The shift to telemedicine has brought about additional challenges for parent physicians delivering clinical care from home. Further study into ophthalmologist work-related stressors and gender is warranted.

Our study supports concerns for increased overall stress to all ophthalmologists as a result of the COVID-19 pandemic. This can have potentially devastating effects within the medical community as high stress levels may lead to the well-known burnout syndrome and negatively impact patient care.^29^ Through our analysis of gender differences amongst academic ophthalmologists in the United States as it pertains to work-life responsibilities and stress both pre- and amidst the COVID-19 pandemic, we encourage future studies to identify strategies for wellness among all ophthalmologists and lessen physician burnout and withdrawal from the field.

## Data Availability

All data produced in the present study are available upon reasonable request to the authors

